# Distinct patterns of microbiota and its function in end-stage liver cirrhosis correlate with antibiotic treatment, intestinal barrier impairment and systemic inflammation

**DOI:** 10.1101/2024.07.27.24311099

**Authors:** Laura Buttler, David A. Velazquez Ramirez, Anja Tiede, Anna M. Conradi, Sabrina Woltemate, Robert Geffers, Birgit Bremer, Vera Spielmann, Julia Kahlhöfer, Anke Kraft, Dirk Schlüter, Markus Cornberg, Heiner Wedemeyer, Christine Falk, Marius Vital, Benjamin Maasoumy

**Affiliations:** Department of Gastroenterology, Hepatology, Infectious Diseases and Endocrinology, Hannover Medical School, Hannover, Germany; Institute for Medical Microbiology and Hospital Epidemiology, Hannover Medical School, Hannover, Germany; German Center for Infection Research (DZIF), Hannover-Braunschweig, Germany; Genome Analytics Research Group, Helmholtz Centre for Infection Research, Braunschweig, Germany; Hannover Medical School, Excellence Cluster RESIST, Hannover, Germany; TWINCORE, Centre for Experimental and Clinical Infection Research, Hannover, Germany; Center for Individualized Infection Medicine (CiiM), Hannover, Germany; Institute of Transplant Immunology, Hannover Medical School, Hannover, Germany

**Author notes:** Authors for correspondence. Postal address: Department of Gastroenterology, Hepatology, Infectious Diseases and Endocrinology, Hannover Medical School. OE5210, Carl-Neuberg-Str. 1, 30625 Hannover, Germany. Postal address: Institute for Medical Microbiology and Hospital Epidemiology, Hannover Medical School. OE5210, Carl-Neuberg-Str. 1, 30625 Hannover, Germany. Equally contributed (Marius Vital, Benjamin Maasoumy).

## Abstract

**Background:** Decompensated liver cirrhosis (dLC) is associated with a dysbalanced microbiome, however, reasons for those observations and resulting consequences for patients are largely unexplored. We aimed to characterize bacterial and fungal components of gut microbiota applying quantitative genome-resolved metagenomics and investigate their relation with gut barrier integrity, inflammation and how this impacts the clinical outcome of dLC patients.

**Methods:** Samples were collected prospectively from 95 consecutive hospitalized dLC patients between 2017 and 2022. Metagenomic shot-gun sequencing coupled to flow-cytometric analyses were performed for qualitative and quantitative insights into gut microbiota on a compositional and functional level. Plasma, CRP, Zonulin and CD163 were measured to investigate host functions. Competing risk analyses were performed to compare cirrhosis-related complications within 90 days.

**Results:** Median baseline MELD was 16 and median age 57.6 years. Patients were clustered into three groups (G1-G3) showing greatly distinct microbial patterns. G1 displayed lowest diversity and highest *Enterococcus* relative abundance (77.97 %), whereas G2 was dominated by *Bifidobacteria* (52.31 %). G3 was most diverse and clustered most closely with HC. Bacterial concentrations in patients were lower compared with HC (median 2.65 × 10^9^ cells/gram stool), especially for G1 (median of 2.65 × 10^9^ cells/gram stool); G2 and G3 were in-between the two. Fungi were primarily detected in patient samples and an overgrowth in G1 that was dominated by Candida spp (51.63 %) was observed. Moreover, G1-patients most frequently received antibiotics (n=33; 86.8 %) at baseline and had higher plasma levels of Zonulin (p=0.044), CD163 (p=0.019) and a numerically higher incidence of infections (p=0.09).

**Conclusion:** Different bacterial clusters were observed at qualitative and quantitative levels and correlated with fungal abundance. Antibiotic treatment contributed to dysbiosis in patients with dLC, which translated into impairment of the intestinal barrier, translocation and systemic inflammation.

## Introduction

Chronic liver diseases are frequently accompanied by changes of the intestinal microbiome.^1^ Alterations can already be detected in early stages of liver disease and become increasingly aggravated with the progression of hepatic fibrosis.^1^ ^2^ Finally, patients with decompensated liver cirrhosis exhibit a disrupted microbiome, characterized by a dramatically increased abundance of potentially pathogenic bacteria and a reduction of commensal microbes.^3^ Particularly, elevated proportions of the pathobionts *Enterococcus*, *Streptococcus* and *Enterobacteriacae* have been reported.^4^ Along with a dysbalanced bacterial composition are altered microbiome functions resulting in decreased levels of bacterial key metabolites, such as short chain fatty acids (SCFA) and secondary bile acids (sBA). SCFA are fermentation end-products with acetate, butyrate and propionate as the main components that are generated by a myriad of diverse taxa that show decreased abundances in cirrhosis.^5^ ^6^ ^7^ ^8^ Physiologically, SCFA feed the epithelium promoting an intact gut barrier and act anti-inflammatory via modulating the immune landscape.^9^ Reduced levels of SCFA-producing bacteria with a concomitant increase of pathobionts impair the intestinal barrier, which may facilitate the translocation of proinflammatory components into the circulation. This may significantly contribute to systemic inflammation in patients with cirrhosis, which is considered to be a key factor for the development of cirrhosis-associated immune dysfunction (CAID), infection susceptibility, hepatic decompensation (e.g. encephalopathy) and acute-on-chronic-liver failure (ACLF).^10^ ^11^ ^12 13^ ^14^ However, causes of intestinal dysbiosis and direct clinical consequences in patients with end-stage liver disease remain largely unexplored as studies including a clinical follow-up remain sparse in this population. Recently, Lehmann and colleagues demonstrated that a lower intestinal diversity with an abundance of either Entercocci or *Enterobacterales* species was linked to a lack of intestinal SCFA and a higher risk for infections in patients with end-stage liver cirrhosis undergoing liver transplantation. Importantly, they also documented that these severe alterations of the gut microbiota were not present in all patients with advanced liver cirrhosis.^15^ A relative *Enterococcus* abundance of greater than 20 % was observed in only 40 %, while in about 25 % of the patients microbiome diversity was not different from healthy individuals.^15^

Reasons for the differences in the microbiota among cirrhotic patients still need to be determined. One of the factors that might significantly interact with the microbiome is concomitant medication. The frequent need for antibiotics to treat or prevent infections in these patients may induce collateral damage on gut microbiota and influence of frequently prescribed drugs on the intestinal microbes needs, hence, to be determined in the setting of end stage liver disease. Furthermore, bacteria are not the only members of the intestinal microbiota. Recently, the role of the mycobiome gained attention in the context of liver disease and reduced bacterial diversity has also been linked to dysbiotic compositions of intestinal fungi.^16^ However, only little is known about the relevance of fungal dysbiosis in advanced cirrhosis and how it influences the clinical course. Mycotic infections, such as spontaneous fungal peritonitis, are rare, but severe complications in patients with end stage liver disease and the role of fungi in the context of inflammation is still largely in the dark. ^17^

Both treatment of complications and their prevention are essential for patients with decompensated liver cirrhosis. This implicates the question whether distinct microbial features can function as a novel tool in risk stratification and as a therapeutic target in clinical management. Therefore, it is essential not only to identify unfavorable microbial features but gain a detailed understanding for their reason and pathophysiological consequences. Our study provides novel quantitative insights into gut microbiota in in the setting of end stage liver disease. We investigated the impact of medication on the interplay between bacterial and fungal components of gut microbiota and how this affects the clinical phenotype of patients with decompensated liver cirrhosis in terms of inflammation and cirrhosis-associated complications.

## Methods

### Study cohort

All patients were recruited from INFEKTA registry, a prospective observational study including consecutive patients with decompensated liver cirrhosis and ascites (German Clinical Trials Registry ID DRKS00010664). Inclusion criteria are evidence of liver cirrhosis, presence of ascites requiring at least one paracentesis and age of 18 or higher. Exclusion criteria were defined as malignant ascites, HIV-infection, congenital immunodeficiencies and presence of any cancer disease, except for hepatocellular carcinoma within the Milan criteria^18^. Furthermore, patients with a history of organ transplantation, except for those with recurrent cirrhosis after a liver transplant, and complete portal vein thrombosis were excluded. For our analysis, we enrolled all of 95 consecutive patients who were treated at Hannover Medical School between 2017 and 2022 and for whom at least one stool sample was available. A number of 19 healthy subjects served as control group (HC).

### Clinical data and endpoints

Dietary questionnaires were available for a number of 86 patients. The date of sample collection was defined as baseline. Liver transplantation (LTx)-free survival and occurrence of cirrhosis-related complications were observed during 90 days of follow-up. The following clinical complications were assessed:

- any infections, as described previously^19^
- acute-on-chronic-liver-failure (ACLF), as described previously ^20^
- overt hepatic encephalopathy (oHE), according to the West Haven criteria ^21^.

### Processing of fecal samples and bioinformatics analyses

DNA was extracted using the ZymoBIOMICS Miniprep Kit (ZYMO, USA) according to the manufacturer’s protocoll. For metagenomic analyses libraries were prepared (Illumina DNA Prep, Illumina, San Diego, United States) and subsequently sequenced on an Illumina NovaSeq 6000 in paired-end mode (2 × 150 bp) as described previously.^22^ Raw reads were quality filtered using Kneaddata (Huttenhower lab; v0.7.2) and subjected to metaPhlan4 to obtain taxonomic composition. For determining pathways of specific key functions, namely, production of the SCFAs butyrate and propionate and of secondary bile acids, gene catalogues from UHGG.v2 representative were used for mapping via BBmap (from JGI; v38.22; paired-end mode) as described previously.^22,23^ Reconstruction of genomes was done using metaWRAP (v1.3.0) as outlined previously using cut-offs of 80 % completeness and 10 % contamination. ^22^ Genomes were annotated based on the genome taxonomy database (gtdb) using gtdb-tk (v2.1.0).^24^

Bacterial load, expressed as bacterial concentration per gram stool, was determined by fluorescent staining combined with flow cytometry as described previously. ^22^

For mycobiome analyses the ITS region was amplified using a two-step approach as outlined in ^25^ and obtained amplicons were sequenced on Illumina MiSeq (2 × 300 bp) as described previously. ^22^ Sequences were processed via the DADA2 pipeline (v1.20) in R (4.2.2) and annotated by blasting merged sequences against the UNTIE database (v29.11.22) where 70 % identity and 70 % query coverage were set as cut-offs. The number of reads assigned as fungal origin were recorded for abundance estimations. For metagenomic based analyses quality filtered and decontaminated reads were mapped to fungal reference sequences using BBmap and relative abundance is expressed as percentage of mapped reads of total reads.

Hierarchical clustering was based on Bray Curtis dissimilarities (BC) using the functions *vegdist* and *hclust* (method=ward.D2) from the vegan package (v2.5.7). Networks were based on Spearman correlations (p<0.01 and Spearman’s rho>0.3) and were visualized via cytoscape (v3.7.2). Phylogenetic trees were constructed from single copy house-keeping genes using gtdb-tk. Detection of *vanA, vanB* and *vanC* genes in samples was done by ariba.

### Measurement of plasma Zonulin and CD163 levels

Plasma zonulin and CD163 levels were measured in a number of 82 patients with available plasma samples as surrogate for intestinal permeability and macrophagge activation following bacterial translocation. Enzyme-linked immunosorbent assays (catalog number abx151842, abbexa, Leiden, NL and DC1630, R&D Systems, Minneapolis, MN) were performed according to the respective manufacturer’s instructions.

### Statistical analysis

Baseline characteristics of patients were analyzed using IBM SPSS Statistics (Version 28, IBM®, New York) and R Statistical Software (version 4.2.0, R foundation for statistical Computing, Vienna, Austria) with the “tableone” package. Categorial values are depicted as number and percentage and compared in a Chi-Square test. Continuous parameters, shown as median and interquartile range (IQR) were analyzed with Man-Whitney-U test or with Kruskal-Wallis-test for comparisons of more than two groups. Correlation of patient parameters with bacterial and fungal microbiota were done in R using regression analyses (function *lm*) and spearman correlation. Boxplots were created with GraphPad Prism (version 10.0). Competing risk analyses were done with R commander and plugin “EZR”.

Competing risk analyses were performed to compare LTx-free survival and the cirrhosis-associated complications (treating LTx or LTx and death as competitors, respectively). Patients were censored with the time point of LTx or end of follow-up. In a first step, the clinical outcome of microbiota groups was compared. Here, G1 functioned as reference group. A number of three patients clustered with HC and were therefore not considered for group comparisons. In a second step, the impact of increased Zonulin (median HC Zonulin level fourfold increased) and elevated CD163 concentrations (median HC CD163 level tenfold increased) on clinical complications was analyzed.

### Ethics

Our study was approved by the ethics committee of Hannover Medical School (Nr. 3188-2016) and respected the declarations of Helsinki. All patients gave permission for the analysis of their data and biomaterial in the form of written informed consent.

## Results

### Clinical characteristics of the overall study cohort

Median age was 57.6 years and 76.1 % were male patients (n=70). Median baseline MELD was 16 (**Table 1a**). Most common cause of cirrhosis was alcohol-related (n=59; 64.1 %). At the time point of stool collection, 8.7 % of the patients (n=8) was diagnosed with SBP, almost a third (n=28; 30.4) had any infection, 14.1 % had ACLF (n=13) and 16.3 % (n=15) showed symptoms of oHE.

**Table 1a.** Baseline characteristics.

|  | All patients<br>(n=95) | Group 1<br>(n=39) | Group 2<br>(n=24) | Group 3<br>(n=29) | p value* |
| --- | --- | --- | --- | --- | --- |
| **Age (y)** | 57.6<br>(50.9-65.5) | 59.41<br>(51.07-65.33) | 56.64<br>(52.98-60.88) | 55.83<br>(50.63-71.73) | 0.715 |
| **Sex** |  |  |  |  | 0.885 |
| - Male | 70 (76.1) | 29 (74.4) | 18 (75.0) | 23 (79.3) |  |
| - Female | 22 (23.9) | 10 (25.6) | 6 (25.0) | 6 (20.7) |  |
| **Etiology\*\*** |  |  |  |  |  |
| - ALD | 45 (48.9) | 18 (46.2) | 14 (58.3) | 13 (44.8) | 0.559 |
| - MetALD | 14 (15.2) | 4 (10.3) | 4 (16.7) | 6 (20.7) | 0.483 |
| - MASLD | 9 (9.8) | 2 (5.1) | 3 (12.5) | 4 (13.8) | 0.430 |
| - Viral | 5 (5.4) | 3 (7.7) | 1 (4.2) | 1 (3.4) | 0.710 |
| - PSC | 4 (4.3) | 1 (2.6) | 2 (8.3) | 1 (3.4) | 0.530 |
| - PBC | 3 (3.3) | 2 (5.1) | 0 (0.0) | 1 (3.4) | 0.537 |
| - AIH | 7 (7.6) | 2 (5.1) | 2 (8.3) | 3 (10.3) | 0.716 |
| - cryptogenic | 10 (10.9) | 6 (15.4) | 1 (4.2) | 3 (10.3) | 0.379 |
| - others | 8 (8.7) | 4 (10.3) | 0 (0.0) | 4 (13.8) | 0.187 |
| **MELD** | 16 (12-22) | 18 (14-24) | 15 (12-20) | 12 (10-14) | 0.004 |
| **SBP at BL** | 8 (8.7) | 3 (7.7) | 1 (4.2) | 4 (13.8) | 0.445 |
| **Infection at BL** | 28 (30.4) | 12 (30.8) | 9 (37.5) | 7 (24.1) | 0.574 |
| **ACLF at BL** | 13 (14.1) | 8 (20.5) | 4 (16.7) | 1 (3.4) | 0.125 |
| **HE at BL** | 15 (16.3) | 7 (17.9) | 4 (16.7) | 4 (13.8) | 0.899 |
| **Laboratory values** |  |  |  |  |  |
| **White blood cells (Tsd/μl)** | 6.00<br>(3.80-8.85) | 6.30<br>(3.65-10.55) | 5.70<br>(3.08-8.18) | 4.60<br>(4.20-8.10) | 0.703 |
| **Red blood cells (Mio/μl)** | 3.00<br>(2.58-3.44) | 2.77<br>(2.46-3.20) | 3.01<br>(2.75-3.42) | 3.20<br>(2.85-3.45) | 0.093 |
| **Hemoglobin (g/dl)** | 9.35<br>(8.23-11.10) | 9.00<br>(7.80-9.80) | 9.35<br>(8.65-10.32) | 10.30<br>(8.50-11.60) | 0.052 |
| **Platelets (Tsd/μl)** | 101.50<br>(61.50-168.75) | 94.00<br>(57.50-145.00) | 85.50<br>(61.25-201.25) | 117.00<br>(88.00-152.00) | 0.310 |
| **INR** | 1.31 (1.16-1.56) | 1.48<br>(1.27-1.62) | 1.38<br>(1.20-1.51) | 1.16<br>(1.10-1.32) | 0.001 |
| **Potassium (mmol/l)** | 4.1 (3.8-4.6) | 4.20<br>(3.85-4.60) | 3.90<br>(3.70-4.32) | 4.30<br>(3.90-4.60) | 0.259 |
| **Sodium (mmol/l)** | 134.50<br>(131.00-138.00) | 134.00<br>(131.00-136.00) | 136.00<br>(132.75-139.00) | 134.00<br>(130.00-137.00) | 0.294 |
| **Bilirubin (μmol/l)** | 27.00<br>(14.00-94.00) | 55.00<br>(18.50-134.50) | 21.00<br>(15.00-46.75) | 17.50<br>(11.75-44.75) | 0.021 |
| **Creatinine (μmol/l)** | 115.00 (90.00-143.00) | 115.00 (96.50-139.00) | 119.00 (88.75-144.75) | 114.50 (80.50-139.50) | 0.815 |
| **AST (U/l)** | 42.00 (32.00-66.75) | 46.00 (33.00-75.50) | 39.50 (33.00-53.00) | 41.00 (32.00-82.00) | 0.683 |
| **ALT (U/l)** | 26.00<br>(18.00-45.00) | 26.50<br>(19.25-44.25) | 25.50<br>(17.75-32.50) | 24.00<br>(17.00-52.00) | 0.960 |
| **AP (U/l)** | 142.50<br>(101.25-201.50) | 134.00 (98.00-168.00) | 152.00 (92.75-220.25) | 162.00<br>(116.00-220.00) | 0.182 |
| **Gamma-GT (U/l)** | 118.00<br>(59.50-221.00) | 90.00<br>(43.50-170.50) | 151.00<br>(73.00-257.50) | 142.00<br>(71.00-259.00) | 0.041 |
| **CRP (mg/l)** | 16.80<br>(8.00-30.00) | 21.40<br>(9.40-32.40) | 11.50<br>(6.05-24.75) | 14.20<br>(8.15-21.65) | 0.183 |
| **Serum-cholinesterase (kU/l)** | 2.04<br>(1.26-2.71) | 1.54<br>(1.18-2.20) | 2.09<br>(1.50-2.96) | 2.21<br>(1.37-3.26) | 0.183 |
| **Albumin (g/l)** | 30.00<br>(25.00-35.00) | 28.00<br>(26.00-31.00) | 33.00<br>(25.00-36.00) | 28.00<br>(27.00-33.00) | 0.467 |
| **Diabetes mellitus** | 24 (26.1) | 7 (17.9) | 8 (33.3) | 9 (31.0) | 0.307 |
| **Active alcohol consumption** | 9 (10.2) | 2 (5.6) | 2 (8.3) | 5 (17.9) | 0.256 |
| **<u>Medication</u>** |  |  |  |  |  |
| **Any antibiotics within one week before BL** | 67 (75.3) | 33 (89.2) | 19 (79.2) | 15 (51.7) | 0.004 |
| **Norfloxacin within 1 week before BL** | 11 (12.2) | 7 (18.9) | 1 (4.2) | 3 (10.3) | 0.213 |
| **Rifaximin within 1 week before BL** | 38 (41.3) | 14 (35.9) | 16 (66.7) | 8 (27.6) | 0.011 |
| **Any other antibiotics** | 39 (42.4) | 28 (71.8) | 4 (16.7) | 7 (24.1) | <0.001 |
| - Vancomycin |  |  |  |  |  |
| - Meropenem | 6 (6.2) | 6 (15.4) | 0 (0.0) | 0 (0.0) | 0.013 |
| - Ampicillin/Sulbactam | 7 (7.6) | 5 (12.8) | 1 (4.2) | 1 (3.4) | 0.269 |
| - Piperacillin/Tazobactam | 7 (7.6) | 5 (12.8) | 1 (4.2) | 1 (3.4) | 0.269 |
| - Ceftriaxone | 10 (10.9) | 8 (20.5) | 1 (4.2) | 1 (3.4) | 0.039 |
| - Clindamycin | 11 (12.0) | 9 (23.1) | 1 (4.2) | 1 (3.4) | 0.019 |
|  | 3 (3.3) | 1 (2.6) | 0 (0.0) | 2 (6.9) | 0.353 |
| **Number of different antibiotics** | 1.00<br>(0.00-2.00) | 2.00<br>(1.00-2.50) | 1.00<br>(0.00-1.00) | 1.00<br>(0.00-1.00) | <0.001 |
| **Non selective beta blockers at BL** | 48 (52.7) | 19 (50.0) | 13 (54.2) | 16 (55.2) | 0.904 |
| **Carvedilol** | 40 (44.0) | 18 (47.4) | 11 (45.8) | 11 (37.9) | 0.726 |
| **Propranolol** | 8 (8.8) | 1 (2.6) | 2 (8.3) | 5 (17.2) | 0.112 |
| **Proton pump inhibitors at BL** | 74 (81.3) | 32 (84.2) | 18 (75.0) | 24 (82.8) | 0.644 |
| **Lactulose at BL** | 57 (62.6) | 23 (60.5) | 19 (79.2) | 15 (51.7) | 0.114 |
| **Ornithine aspartate at BL** | 28 (30.8) | 8 (21.1) | 12 (50.0) | 8 (27.6) | 0.050 |

Values as number (percentage) or median (interquartile range). \* Chi-Square was used for comparison of categorical variables, Fisher-Freeman-Halton's exact for categorial covariables with less than five cases/field. Kruskal-Wallis test was performed for continuous values.
\*\*Results in more than 100 % due to mixed etiology.

### Gut bacteria clustered into three compositionally distinct groups that varied in concentrations and in abundances of key functions

Diversity and composition of gut bacteria varied widely between patients and hierarchical clustering suggested three groups that were separated form healthy controls (HC). Group one (G1) was dominated by *Enterococcus sp* (77.97 % (51.67-95.44)), whereas *Bifidobacteria* were most abundant in group two (G2) (52.31 % (42.82-65.52)) (**Figure 1A; Suppl. figure 1**). The third group (G3) clustered most closely with HC showed a diverse pattern. Diversity based on observed species and the Shannon index was greatly distinct between groups, where G1 showed lowest values of 10 (8-17) (observed species) and 0.74 (0.22-1.37) (Shannon) followed by G2 (56 (42-79); 2.17 (1.98-2.62)) and G3 (62 (44-87); 2.72 (2.23-3.02)). HC displayed highest diversities with 261 (200 - 313) species and a Shannon index of 3.94 (3.68-4.28) (**Figure 1A; Suppl. figure 1**). Bacterial concentrations determined by flow cytometry followed this pattern: G1 was characterized by a low bacterial load of 2.65 × 10^9^ cells/gram stool (1.21 × 10^9^-6.67 × 10^9^) , which was 2 factors of magnitude lower compared with HC (2.38 × 10^11^ cells/gram stool (1.71 × 10^9^-3.16 × 10^9^). G2 and G3 showed higher values of 2.55 × 10^10^ cells/gram stool (5.52 × 10^9^-3.77 × 10^10^) and 1.52 × 10^10^ cells/gram stool (7.40 × 10^9^-2.65 × 10^10^), respectively, that were, however, still markedly below HC. Similar were results of abundances of microbial key pathways encoding enzymes for the synthesis of the SCFA butyrate and propionate as well as for secondary bile acids (sBA) (**Figure 1A**). Genes associated with butyrate and sBA were almost absent in G1 and also lower in G2 (6.47 % (1.06-10.59) and 0.00 % (0.00-0.20)) and G3 (13.02 % (2.27-24.29) and 0.03 % (0.00-0.71)) compared with HC, where 31.49 % (26.57-33.49) and 0.54 % (0.33-1.21), respectively, of calculated genomes harbored genes of those pathways (**Figure 1A; Suppl. figure 2**). Propionate pathways had higher values in patient groups G2 (pdiol: 5.82 % (2.47-14.26), suc: 8.66 % (2.94-16.48)) and G3 (pdiol: 10.49 % (5.30-26.31), suc: 13.02 % (2.27-24.29)) displaying similar concentrations as those observed in HC (pdiol: 7.04 (5.03-8.69), suc: 12.12 (6.53-16.57)); with a few exceptions those pathways were lower in samples derived from G1.

**Figure 1.**
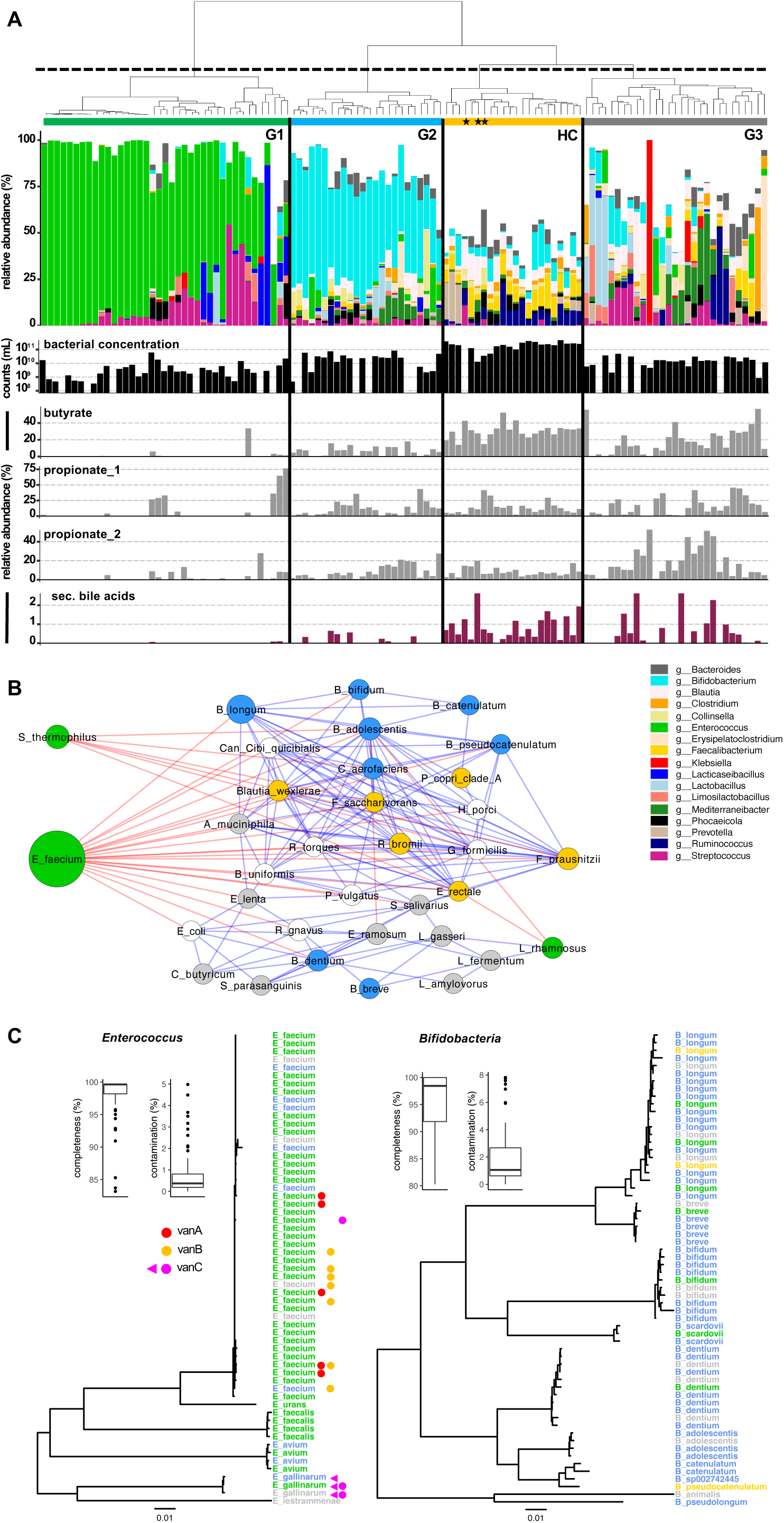
Bacterial communities of patients and healthy controls (HC). Panel **A** shows hierarchical clustering result along with composition on the genus level for all samples yielding three distinct patient groups (G1-G3) and a separate HC group (the black stars mark samples from patients that grouped together with HC). Below bacterial cell concentrations measured by flow cytometry (bacterial concentration) as well as relative abundances of pathways for the synthesis of butyrate, propionate (two pathways) and secondary bile acids are given. In panel **B** network of bacterial species based on correlation analysis is given, where node size refers to mean abundance of each species and colors of edges represent positive (blue) and negative (red) correlations. Species primarily associated with a specific group from hierarchical clustering are color coded. Panel **C** shows phylogenetic trees for genomes of *Enterococcus* and *Bifidobacteria* assembled from metagenomes of patient samples along with their basic quality parameters completeness and contamination. Group origins of genomes are indicated by color and presence of genes encoding vancomycin resistance (*vanABC)* in respective samples is given; the violet triangle signifies *vanC* detection on genomes.

Correlation analyses revealed separate clustering of signature taxa of individual groups. *E. faecium*, which was the main *Enterococcus* species, was negatively associated with a variety of other taxa such as G2’s *Bifidobacteria* species that, apart from *B. breve* and *B. dentium*, clustered closely together (**Figure 1B**). Other taxa specific for G1, namely, *Streptococcus thermophilus* also correlated negatively with several taxa of G2 and HC. *Faecalibacterium prausnitzii* of HC formed a counterpart that was positively associated with a variety of taxa of HC as well as of G2 and G3, whose nodes formed a separate group.

In total, we assembled 645 high-quality genomes of patient samples, where 59 and 62 genomes were annotated as *Enterococcus* and *Bifidobacteria*, respectively (**Figure 1C**). The former was primarily derived from samples of G1, where genomes were retrieved from 37 (94.9 %) samples and the bulk was annotated as *E. faecium*. In five samples two genomes of this taxon were obtained (one sample harbored three *Enterococci* genomes) with an average completeness of 97.79 % ± 3.84 % and a contamination of 0.84 % ± 1.17 %. Genes associated with Vancomycin resistance were detected in 14 samples (11 (38 %) in G1), however, apart from vanC in all three *E. gallinarum* strains *E. faecium* genomes were suspiciously devoid of *vanA,B* genes, most probably due to their presence on plasmids. For *Bifidobacteria*, genomes from several distinct species were assembled that primarily derived from samples of G2. Their average completeness was 95.59 % ± 5.55 % with a contamination of 2.07 % ± 2.21 % and up to four genomes were detected in one sample.

### Abundances of fungi were greatly distinct between patient groups and correlated with specific bacterial taxa

We analyzed fungal organisms based on three methods, namely, (i) amplification and sequencing of the ITS region and by two metagenome-based analyses comprising (ii) a custom workflow based on mapping reads to a comprehensive fungal database comprising a multitude of fungal genes and (iii) MetaPhlan4 that uses specific single copy marker genes. Results between ITS and the custom metagenomic approach were largely congruent and detected much higher fungal abundances in G1 compared with other groups (**Figure 2A; Suppl. figure 1**). Samples of HC were largely devoid of fungal communities. Four genera dominated fungal compositions with species belonging to *Candida spp* being the most abundant ((51.63 % (6.53-95.45)). Fungi of the related genus *Nakaseomyces* (0.00 % (0.00- 13.24)) and of the family *Saccharomycetaceae, Saccharomyces* (0.12 % (0.0-7.00)) and *Kluyveromyces* (0.00 % (0.00-0.01)) were detected in patients as well, where results of ITS and the custom bioinformatic approach yielded congruent insights (**Figure 2A**). Results based on MetaPlan4 were supporting the general trend of G1 being most colonized by fungi, however, the method could only detect taxa of the genera *Candida* and *Saccharomyces,* and showed lower sensitivities.

**Figure 2.**
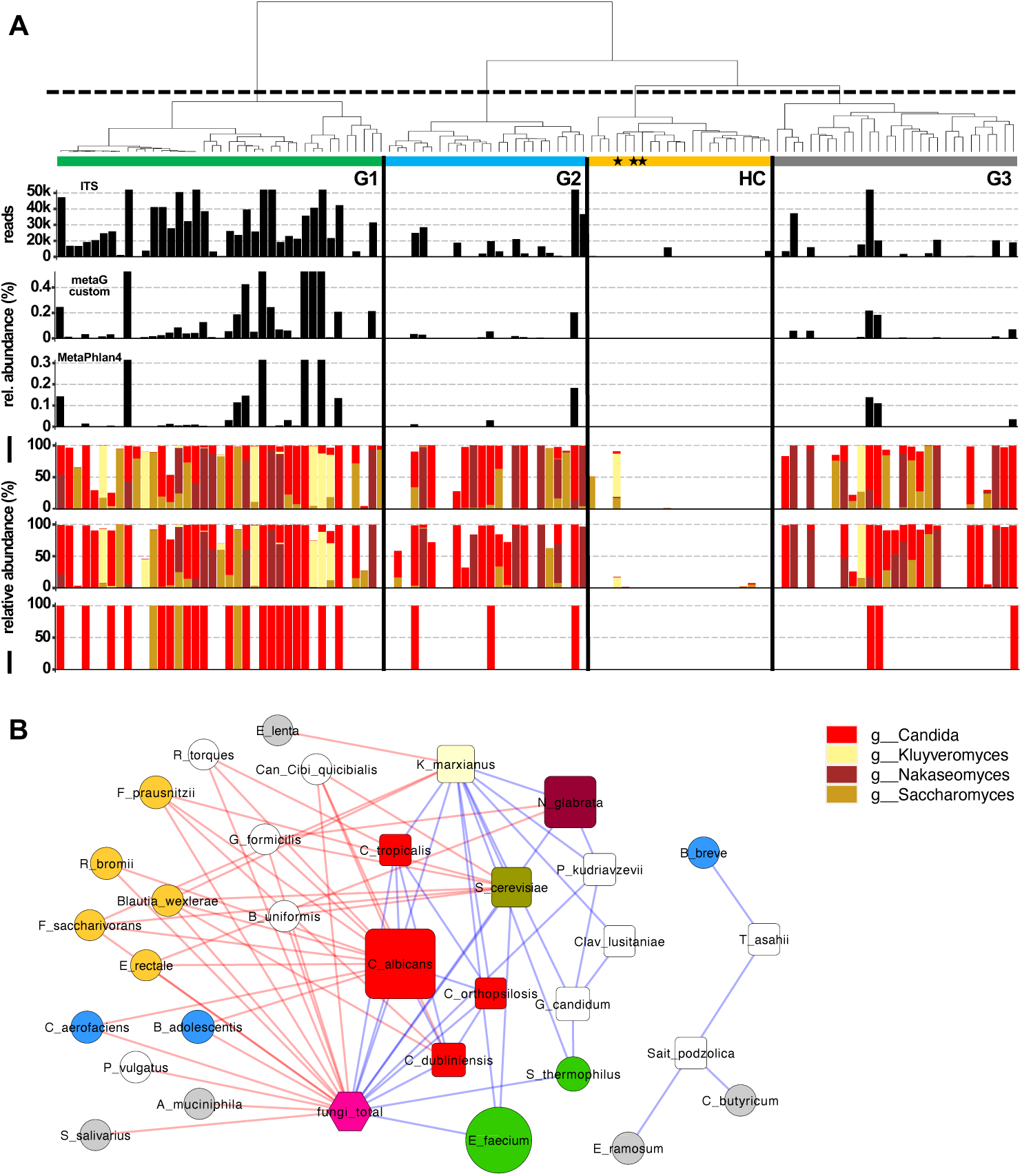
Fungal communities of patients and healthy controls (HC). Results in panel **A** are displayed according to groupings based on hierarchical clustering result of bacterial communities that are shown in Figure 1. Three different methods were applied based on (i) amplification of ITS region, (ii) custom metagenomic analysis and (iii) MetaPhlan4. Fungal concentrations measured by reads associated with fungi derived from the individual methods are given as black bars, whereas composition on the genus level is given in the three barplots below (same order as above). In panel **B** a network of fungal species and associated bacterial species based on correlation analyses is given, where node size refers to mean abundance of each species and colors of edges represent positive (blue) and negative (red) correlations. Bacterial species primarily associated with a specific group from hierarchical clustering are color coded (compare Figure 1).

Correlation analyses between bacterial and fungal species revealed a clear pattern, where total concentrations of fungi were positively correlated only with bacterial taxa of G1 (*E. faecium* and *S. thermophilus*), whereas bacteria of samples from other patient groups were all negatively correlated with total fungi and individual fungal species. A few species (n=5) formed a separate module (**Figure 2B**).

### Antibiotic treatment significantly contributes to distinct intestinal microbiota compositions

No differences in age and cirrhosis etiology was detected between groups, whereas median MELD was higher in G1 than in the other groups. Administration of frequently prescribed drugs, e.g. non selective beta blockers and proton pump inhibitors was almost equally distributed over the patient cohort (**Table 1a**). Furthermore, dietary and lifestyle habits were comparable between groups (**Table 1b**).

**Table 1b.** Diet and lifestyle of the study cohort.

|  | All patients<br>(n=86) | Group 1 (n=35) | Group 2 (n=24) | Group 3 (n=27) | p value* |
| --- | --- | --- | --- | --- | --- |
| Coffee<br>drinkers | 57 (66.3) | 25 (71.4) | 13 (54.2) | 19 (70.4) | 0.334 |
| Smokers | 31 (32.0) | 12 (32.4) | 11 (45.8) | 8 (29.6) | 0.432 |
| Vegetarian | 5 (5.8) | 2 (6.1) | 1 (4.2) | 2 (7.4) | 0.887 |
| > 2 fruits/day | 22 (26.5) | 7 (21.9) | 7 (29.2) | 8 (29.6) | 0.750 |
| vegetable<br>intake ≥ 5<br>days/week | 45 (60.8) | 20 (66.7) | 10 (50.0) | 15 (62.5) | 0.486 |

A high proportion of 75.3 % (n=67) received antibiotic treatment within one week before stool was provided (**Table 1a**). The number of patients with intake of any antibiotics differed significantly between groups (p=0.004), where G1 contained the highest quantity of patients with antibiotic treatment (n=33; 86.8 %), followed by G2 (n=19; 79.2 %). Furthermore, the median number of different antibiotics per patient was significantly higher in G1 (p<0.001). In addition, the type of administered antibiotics differed. Besides a significantly higher intake of Ceftriaxone in patients from G1 (p=0.019), also the administration of Vancomycin (p=0.013) and Piperacillin/Tazobactam (p=0.039) was higher in this group, whereas antibiotic treatment in G2 was mainly dominated by Rifaximin (n=16; 66.7 %; p=0.011) (**Table 1a**).

### Abundance of Enterococcus is linked to an increase in surrogate markers of an impaired intestinal barriers and bacterial translocation

Significantly greater concentrations of Zonulin were discovered in cirrhosis patients compared to HC (9.5 ng/ml (6.4-16.3) vs. 4.2 ng/ml (2.8-8.7); p=0.023). Patients of G1 reached highest median Zonulin values that exceeded the measured levels of the other patients significantly (12.8 ng/ml (8.0-17.6) vs. 8.2 ng/ml (5.2-13.6); p=0.044) (**Figure 3A, B**). Similarly, CD163 concentrations were significantly elevated in LC patients contrasted to healthy subjects (2193.0 ng/ml (1377.0-3290.8) vs. 388.0 ng/ml (359.5-507.5); p<0.001). Likewise, G1 exhibited significantly higher CD163 levels than the other patients (2544.9 ng/ml (1448.4-4151.4) vs. 1836.0 ng/ml (1356.6-2565.3); p=0.019) (**Figure 3 C, D**).

**Figure 3.**
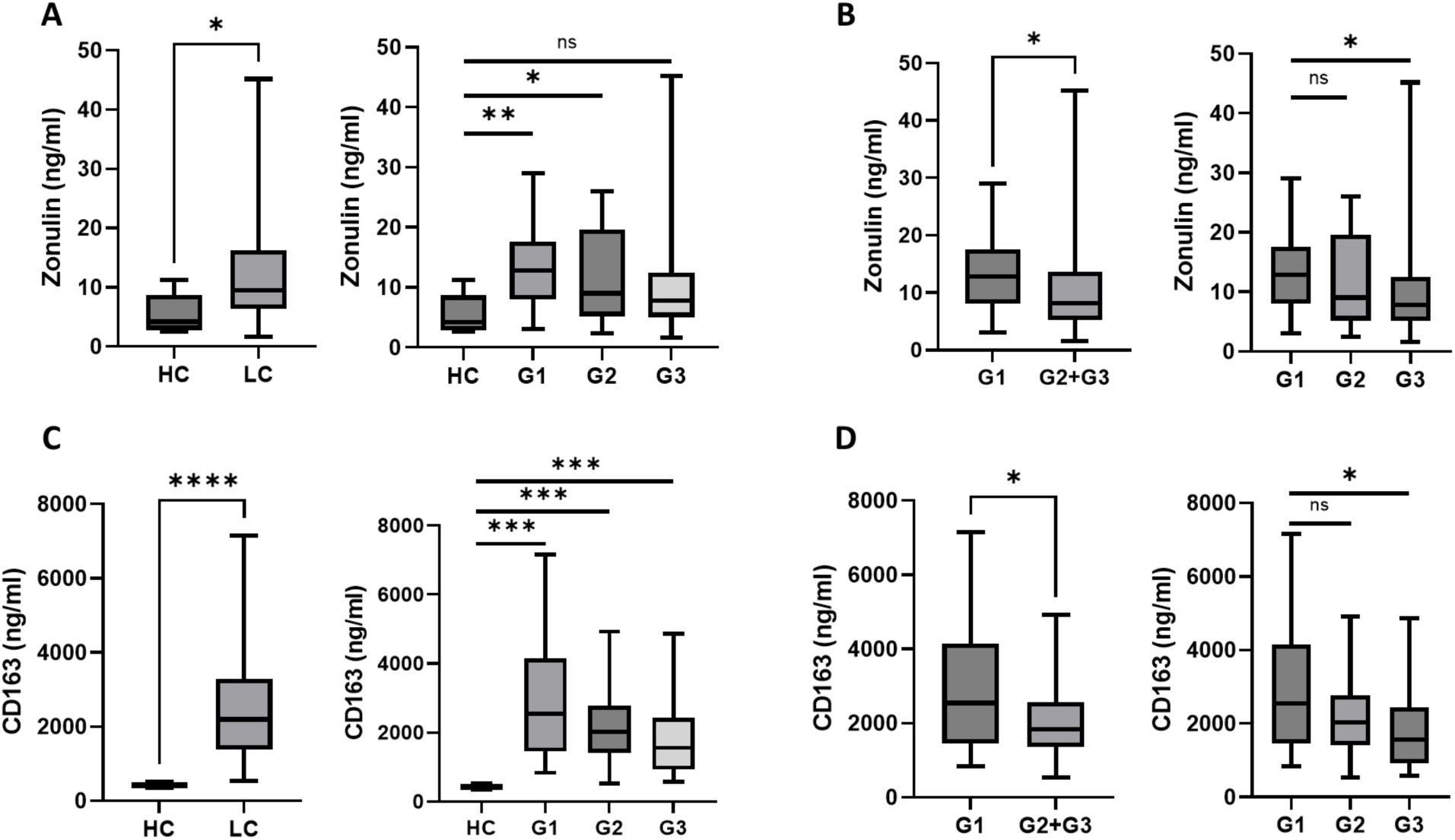
Zonulin levels were higher in patients than in HC (p=0.023) (**A**). Among cirrhosis patients, G1 displayed significantly higher Zonulin concentrations than the other patients (p=0.044) (**B**). CD163 concentrations in cirrhosis patients exceeded those of HC (p<0.001) (**C**), with highest values in G1 compared to the other patients (p=0.019) (**D**).

### Microbiota profiles, impaired intestinal barrier and increased translocation translate in a numerical increase of certain cirrhosis-associated complications

In the overall study cohort, 27.2 % (n=25) of the patients died or underwent LTx within 90 days (**Table 2a**). Almost a third acquired any infection 30.4 % (n=28) with spontaneous bacterial peritonitis (SBP) being the most frequent (n=13; 46.4 %) (**Table 2b**). Mycotic infections were detected in 6.5 % (n=6) subjects within the observational period, with candida esophagitis in 3 cases, UTI in 2 patients and peritonitis in one case. Moreover, an episode of oHE occurred in 21.7 % (n=20) patients and 30.4 % (n=28) developed ACLF.

**Table 2a.** Incidences of complications during 90 days of follow-up.

|  | All patients (n=92) | Group 1 (n=39) | Group 2 (n=24) | Group 3 (n=29) |
| --- | --- | --- | --- | --- |
| **Death/LTx** | 25 (27.2) | 11 (28.2) | 7 (29.2) | 7 (24.1) |
| **Death** | 16 (17.4) | 5 (12.8) | 4 (16.7) | 7 (24.1) |
| **LTx** | 9 (9.8) | 6 (15.4) | 3 (12.5) | 0 (0.0) |
| **Infection** | 28 (30.4) | 14 (35.9) | 8 (33.3) | 6 (20.7) |
| **SBP** | 15 (16.3) | 8 (20.5) | 4 (16.7) | 3 (10.3) |
| **ACLF** | 28 (30.4) | 12 (30.8) | 7 (29.2) | 9 (31.0) |
| **Severe ACLF (≥ grade 2)** | 12 (13.0) | 6 (15.4) | 4 (16.7) | 2 (6.9) |
| **HE** | 20 (21.7) | 9 (23.1) | 4 (16.7) | 7 (24.1) |

Values as number (percentage).

**Table 2b.**
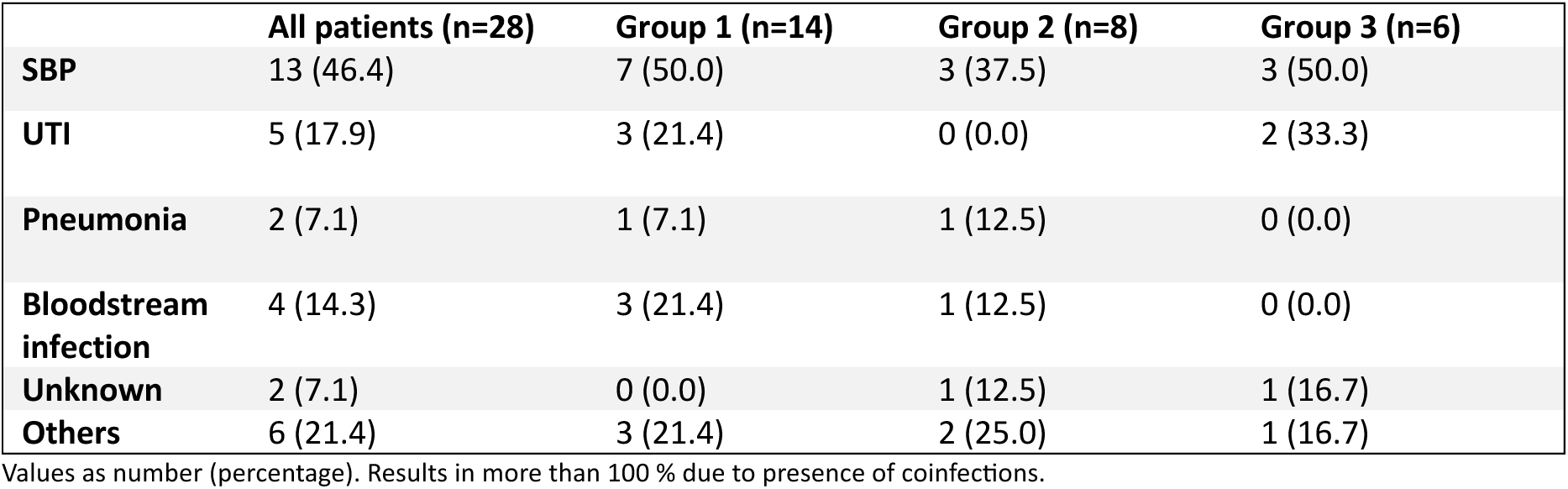
Types of infections during 90 days of follow-up.

**Table 2c.** Infections causing pathogens.

|  | All patients (n=13) | Group 1 (n=8) | Group 2 (n=4) | Group 3 (n=2) |
| --- | --- | --- | --- | --- |
| Enterococcus | 5 (38.5) | 3 (37.5) | 2 (50.0) | 0 (0.0) |
| S. aureus | 2 (15.4) | 1 (12.5) | 1 (25.0) | 0 (0.0) |
| Coagulase negative staphylococcus | 3 (23.1) | 1 (12.5) | 2 (50.0) | 0 (0.0) |
| Candida sp | 6 (46.2) | 5 (62.5) | 0 (0.0) | 1 (50.0) |
| others | 2 (1.4) | 0 (0.0) | 1 (25.0) | 1 (50.0) |

Values as number (percentage). Results in more than 100 % due to presence of coinfections.

Regarding LTx-free survival, no differences between groups were detected in the competing risk analyses, treating G1 as reference group (G2=HR=1.08, p=0.91; G3: HR=1.80; p=0.31) (**Figure 4 A**). Of note, incidences of infections were numerically higher in G1 than in G3 (n=14; 35.9 % vs. n=6, 20.7 %; HR=0.45, p=0.09) (**Table 2a**, **Figure 4 B**). Additionally, the risk for fungal infections was significantly increased among G1 patients compared to G3 (p<0.001) (**Figure 4 C**). Contrastingly, the likelihood for oHE (G2: HR=0.65, p=0.48; G3: HR=1.00, p=1.00) and ACLF (G2: HR=0.78, p=0.61; G3: HR=0.91, p=0.83) was almost equal between groups (**Figure 4 D, E**).

**Figure 4.**
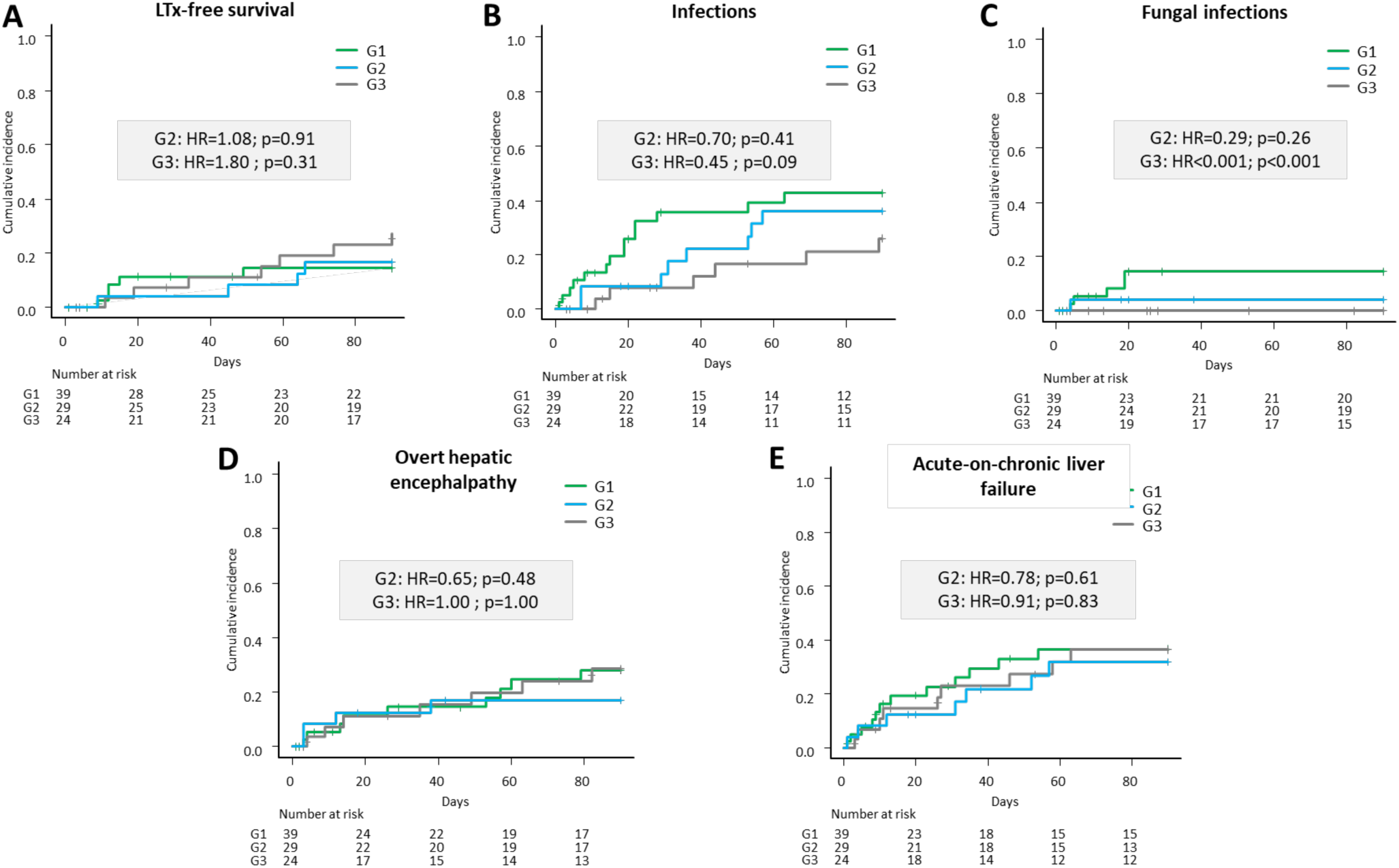
Competing risk analyses of liver transplantation (LTx)-free survival (**A**), infections (**B**), fungal infections (**C**), overt hepatic encephalopathy (**D**) (oHE) and acute-on-chronic liver failure (ACLF) (**E**). Group 1 (G1) was treated as reference group.

In a second step, the impact of increased Zonulin and elevated CD163 levels on the disease course were investigated. Although increased Zonulin concentrations were neither linked to inferior survival (HR=0.81, p=0.75) nor to elevated risk for ACLF (HR=1.28, p=0.60), numerically higher incidences of infectious complications (HR=1.83, p=0.16) and oHE (HR=2.23, p=0.11) were observed in these patients (**Suppl. figure 3**). In line with this, the likelihood for oHE was statistically significantly increased in subjects with elevated CD163 levels (HR=3.27, p=0.02) (**Suppl. Figure 4**).

### Proposal of an integrated model explaining interactions between antibiotic treatment, distinct intestinal microbiota profiles and functioning, intestinal barriers, bacterial translocation and infections in patients with cirrhosis

In order to investigate how different parameters of various levels are connected correlation analyses were performed and a mechanistic model is proposed in **Figure 5**. According to our model antibiotic treatment greatly affected microbiota decreasing bacterial diversity and enriching for *Enterococci*. Concurrently, lower bacterial cell concentrations and increased fungal colonization were observed in those samples. Samples high in *Bifidobacteria* showed opposite patterns. Bacterial key functions, namely, the capacity to synthesize SCFA and sBA, were associated with bacterial concentrations and diversity as well as with *Bifidobacteria* (not sBA), whereas they were negatively associated with *Enterococcus* and fungi. *Enterococci* and lower functional capacities of key metabolites were positively associated with gut permeability (Zonulin) and inflammation (CD163 and CRP) that were also correlated with each other. Elevated abundances of fungi were linked to CRP and the development of infections; antibiotic treatment was associated with infections as well. *Bifidobacteria* did not correlate with any patient parameters (**Figure 5**).

**Figure 5.**
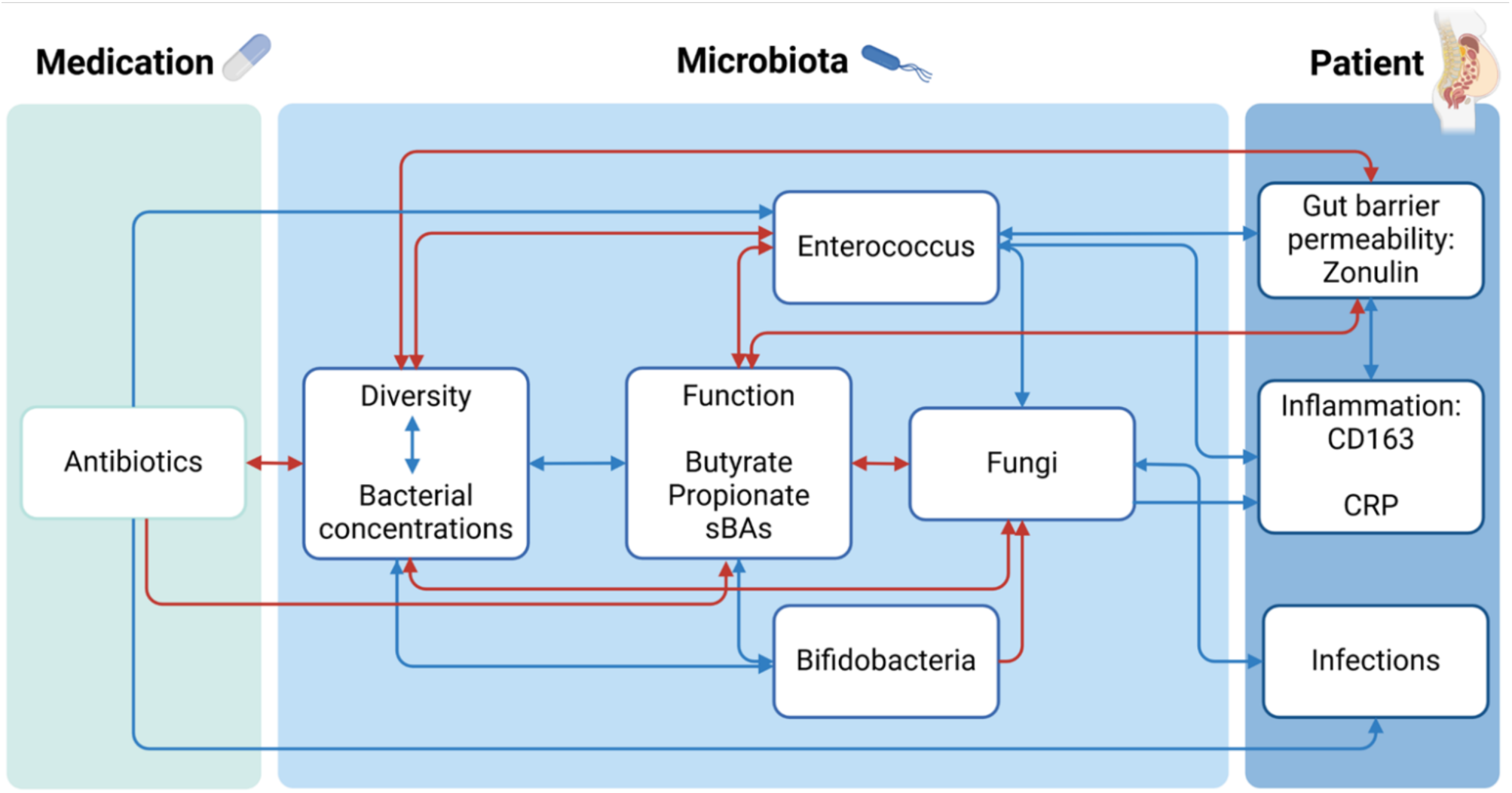
Proposal of an integrated model linking observed results of medication, gut microbiota and clinical characteristics patients.

## Discussion

Liver cirrhosis is characterized by irreversible liver damage that is related to high morbidity and poor survival and accompanied by a dysbalanced intestinal microbiota, where the extent proportionally increases with liver disease stages reaching its peak in decompensated cirrhosis.^26^ ^27^ ^28^ Our study confirmed these prior data and provided additional quantitative insights demonstrating that, next to composition, bacterial load was distinct between patient groups and lower compared to HC. Moreover, we identified medication, specifically antibiotic treatment, as the main parameter explaining the distinct microbial clusters observed, where a particular type dominated by *Enterococcus* was associated with barrier dysfunction and inflammation. Importantly, we integrated a major, but so far largely neglected component of gut microbiota, namely, the mycobiome, discovering an interplay between fungi and specific bacterial taxa that correlated with systemic inflammation and infectious complications.

The gut and the liver are tightly connected via reciprocal organ crosstalk forming the so-called gut-liver axis.^29^ The progression of hepatic disease is, hence, not limited to the liver, but directly influences the intestinal environment and its microbial inhabitants. Typically, a reduction of alpha-diversity that is often associated with increased abundances of pathobionts and lower levels of commensal bacteria have been observed.^30^ Our study supports those results and reiterated a clustering of samples into groups either dominated by *Enterococcus* or *Bifidobacteria* as observed before.^1^ ^15^ ^26^ Those signature taxa correlated with patient parameters, where only the former was correlated with barrier malfunctioning and inflammation. Our study, hence, supports the general view that *Bifidobacteria* are beneficial, also in the context of liver-disease patients, and concentrations of this taxon were associated with higher bacterial loads, higher diversity and a lower fungal burden. Antibiotic treatment was revealed as a major contributor for *Enterococcus* abundance (discussed in detail below), while lactulose, which was recently shown to promote *Bifidobacteria* growth, did not significantly correlate with abundances of this taxon in our study (p=0.18; data not shown). ^26^ Patients associated with G3 that represented the group most closely related to HC displaying highest diversities performed best in many parameters and showed most discrimination to G1 including significantly lower levels of Zonulin and CD163 along with a lower incidence of infections (p<0.1). Those observations strengthen the common view that a microbiota resembling healthy individuals the closest are the most beneficial, also in the context of liver disease, and manipulations thereof should, hence, be done with care as outlined below.

The loss of symbiotic microbes often implicates impaired microbiota functions^15^. Abundances of pathways related to SCFA synthesis and bile acid transformations were markedly lower in our patient cohort compared with HC, which was particularly pronounced in G1. This observation was recently confirmed on a metabolomic level, where microbiota linked to an *Enterococci* bloom were largely devoid of those metabolites.^15^ SCFA serve as nutrition for colonocytes and are, hence, essential for the maintenance the intestinal barrier and act anti-inflammatory. ^5^ ^9^ Thus, reduced SCFA synthesis directly contribute to barrier dysfunction and increased inflammation as indicated by results of our study. To investigate the impact of microbiota patterns on host barrier functions, plasma Zonulin was measured. Zonulin, a haptoglobin 2 pre-protein, is secreted by intestinal epithelial cells in response to stress inducing factors, mainly due to increased pathobiont abundances.^31^ In line with this, cirrhosis patients exhibited significantly increased Zonulin levels compared to HC in our study. Furthermore, Zonulin correlated positively with *Enterococcus* abundance and negatively with microbiota diversity and SCFA production potential, so that Zonulin levels of G1 exceeded those of the other patients markedly. After secretion, Zonulin upregulates tight junction permeability. ^32^ ^33^ Reduced tightness of these intercellular connections allows entrance of gut-derived bacteria and pathogen-associated molecular patterns (PAMPs).^34^ Once PAMPs reach the liver via the portal venous system, liver-resident macrophages (Kupffer cells) quickly respond with the release of CD163.^35^ ^36^ Importantly, the positive correlation between Zonulin and CD163 in our study exemplifies the relationship between increased intestinal permeability, translocation of bacterial products and inflammatory response (CRP). Recently, increased pathobiont abundance has been discussed as predictor for nosocomial infection development in cirrhosis.^37^ Furthermore, expansion of Enterococci over time has been linked to a higher infection vulnerability among patients undergoing LTx.^15^ Our data demonstrate that altered barrier functions might be one out of several contributing factors for this association. In line, G1 patients showed highest incidences of infections during follow-up. However, although incidences differed numerically in our cohort, these disparities were not statistically significant in the competing risk models. This might be explained by a to small sample size. Thus, larger studies will be required to draw final conclusions on this issue.

While some studies have investigated intestinal bacterial composition, our study provides detailed insights on the intestinal mycobiome in patients with decompensated cirrhosis. We demonstrated that the extent of bacterial dysbiosis, along with a reduction of cell concentrations, are related to fungal overgrowth in cirrhosis patients. Importantly, the group with the highest intestinal concentrations of fungi indeed had a higher risk for fungal infections. This is of particular clinical importance as mycotic infections, e.g. spontaneous fungal peritonitis, are severe complications, that potentially cause critical illness and end in up to 30-50 % fatality.^17^ ^38^ Furthermore, fungal abundance did correlate with both inflammatory markers, i.e., CD163 and CRP suggesting a proinflammatory role that potentially contributes to progression of tissue damage, organ dysfunction and therefore to several complications apart from infections in these patients^39^ ^40^. *Enterococcus* was positively associated with fungi as well, however, whether this observation is a result of an interplay between those taxa or confounded by other factors, such as medication, needs to be elucidated in follow-up studies.

To uncover potential causes of the different patterns in our cohort, antibiotic treatment was analyzed. Interestingly, it was found that patients with the lowest cell counts and the highest *Enterococcus* and fungi abundance frequently received broad spectrum antibiotics, which might explain the reduction of autochthonous bacteria. Furthermore, some of these preparations, for example meropenem or ceftriaxone, have relatively low or even no efficacy against Enterococci. This might result in selective growth advantage for this genus. Additionally, these patients received a significantly higher number of different antibiotics within one week before stool was provided. This might further contribute to the diminished autochthonous microbiota and the overall reduction of bacterial cell counts. With the decrease of bacterial concentrations, not only the competition for nutrition, but also the diminished bacterial release of antifungal metabolites, might enable fungi expansion.^41^ ^42^ ^43^ Moreover, beta-lactam antibiotics, that were frequently prescribed to G1 patients, have been linked to the release of peptidoglycans from bacteria.^44^ A recent study reported enhanced growth of *Candida* in the presence of these substances.^44^ Hence, antibiotic treatment seems to contribute to increased fungi abundances via multiple mechanisms. Contrastingly, antibiotic treatment in G2 was mainly dominated by rifaximin that has been linked to a reduction of several pathogenic taxa whilst increasing commensals, such as *Bifidobacteria* and *Faecalibacterium*.^45^ ^46^ ^47^

Once any infection occurs, it will be repeatedly treated with antibiotics.^48^ From the viewpoint of dysbiosis, this might result in a vicious cycle, with repetitive damage of the intestinal microbiota, leading to barrier dysfunction and recurrent infections. Hence, indication for antibiotic treatment should be provided carefully in the setting of advanced liver disease. As the missing longitudinal sample collection is a major limitation of our study, the regenerative capacity of the gut barrier in cirrhosis patients needs to be investigated by further. In contrast, among patients with CHILD C cirrhosis and low ascites protein levels prophylactic treatment with norfloxacin can reduce incidence of SBP and improve one year survival.^49^ ^50^ The preventive effect against SBP might be due to the reduction of gram-negative bacteria, e.g. *Proteobacteria*, that has been observed in rats as well as in cirrhosis patients who received norfloxacin as a prophylaxis. Contrastingly, the abundances of gram-positive cocci remained stable during treatment.^51^ ^52^ However, detailed insights into the microbiota modulating effects of norfloxacin and its impact on long-term changes of the intestinal barrier remain to be determined. Of note, we did not find any significant correlation of norfloxacine prophylaxis with microbiota composition in our study.

In conclusion, distinct individual features of microbiota are present even in the end stage of liver disease and linked to antibiotic treatment. These microbial patterns are strongly associated to intestinal barrier functions that might play an important role for development of cirrhosis-related complications. Further research is required to design novel, microbiota-based treatment options for patients with decompensated liver cirrhosis.

## Data Availability

All data produced in the present study are available upon reasonable request to the authors

## Supplementary figures

**Supplementary figure 1.**
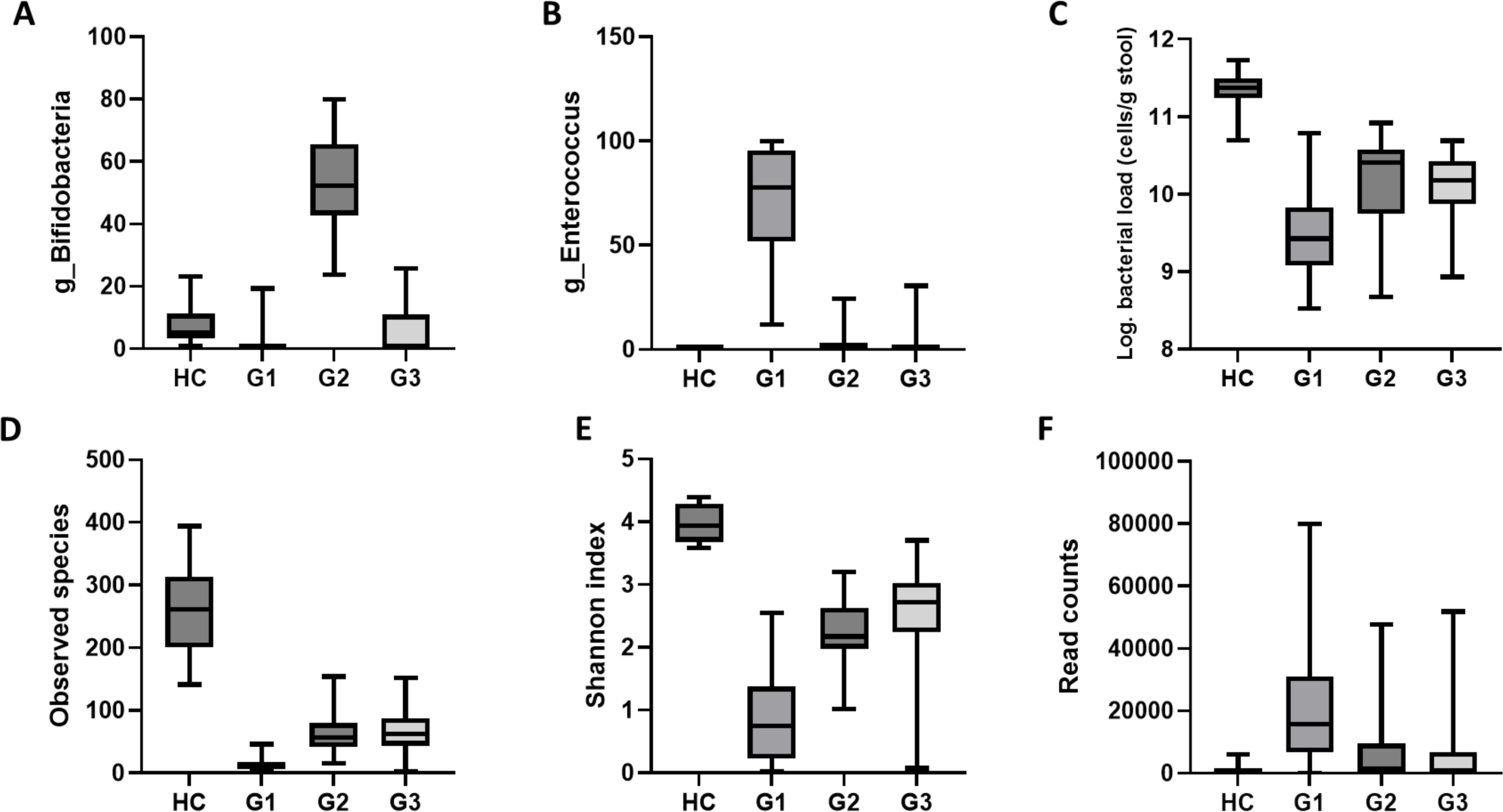
Bifidobacteria were most abundant in G2 (**A**), whereas Enterococcus dominated G1 (**B**). Overall bacterial concentrations were decreased in patients, with lowest values in G1 (**C**). Diversity, assessed by observed species (**D**) and Shannon index (**E**), increased from G1 over G2 and G3 to healthy controls (HC). Fungi are largely devoid in HC, but reached high abundances in G1 (**F**).

**Supplementary figure 2.**
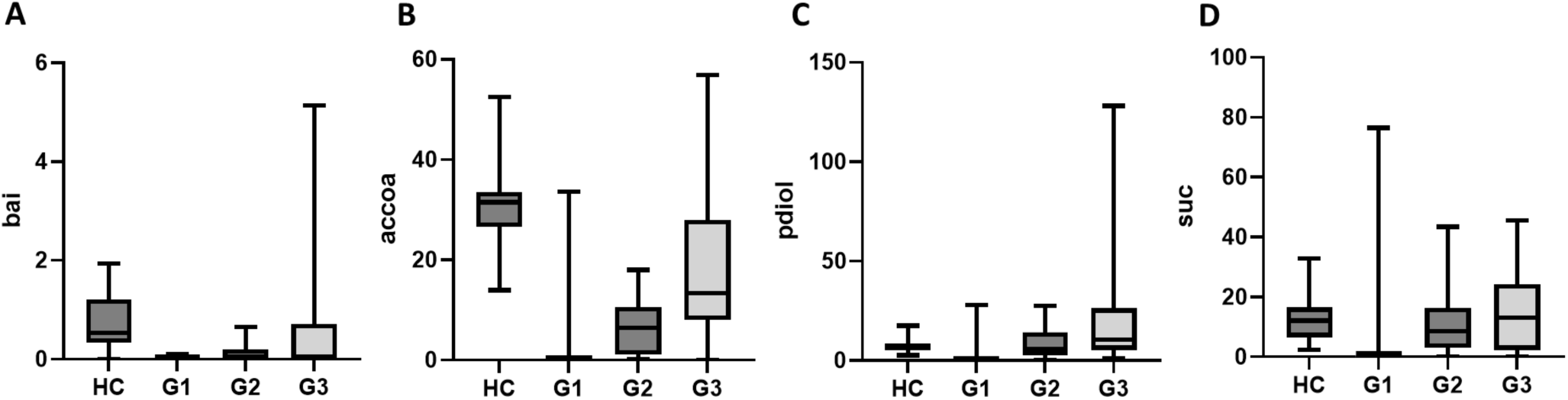
Microbial key pathways encoding enzymes for the synthesis of secondary bile acids (**A**) and the short chain fatty acids butyrate (**B**) and propionate (**C, D**).

**Supplementary figure 3.**
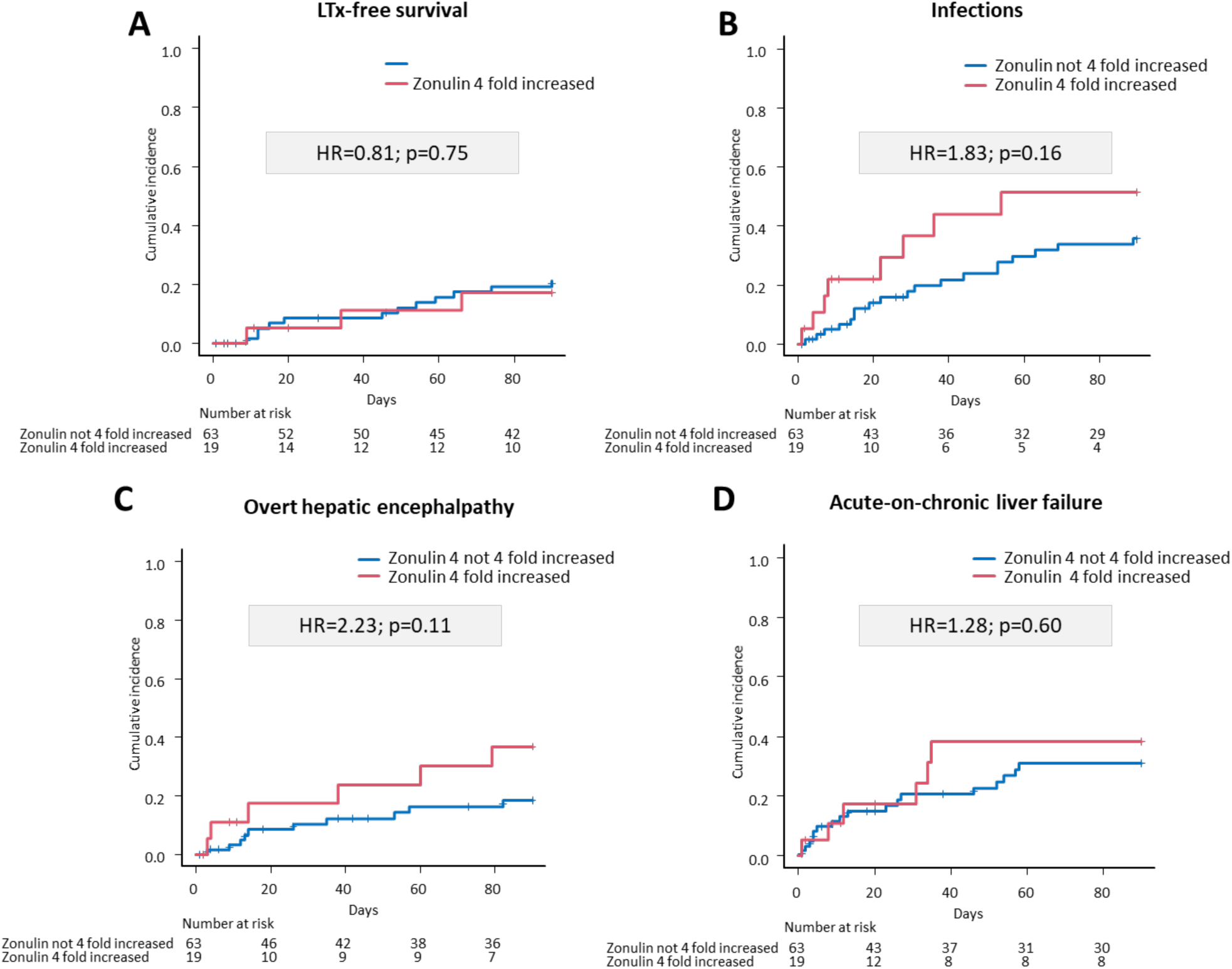
Competing risk analyses of liver transplantation (LTx)-free survival (**A**), infections (**B**), overt hepatic encephalopathy (**C**) (oHE) and acute-on-chronic liver failure (ACLF) (**D**) in patients with and without increased Zonulin levels.

**Supplementary figure 4.**
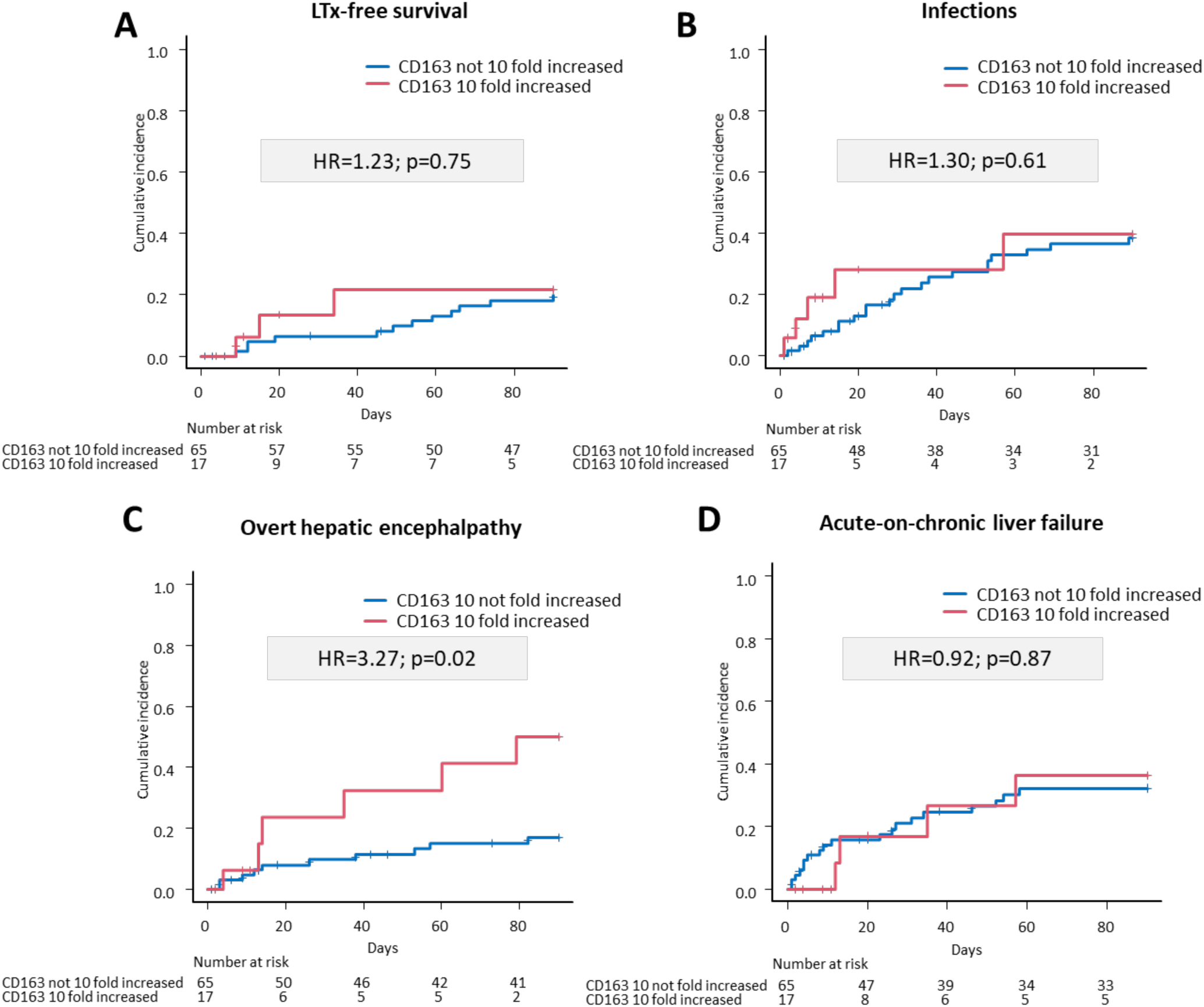
Competing risk analyses of liver transplantation (LTx)-free survival (**A**), infections (**B**), overt hepatic encephalopathy (**C**) (oHE) and acute-on-chronic liver failure (ACLF) (**D**) in patients with and without increased CD163 levels.

